# A consensus diabetes core dataset for research using NHS data: outputs from a Diabetes Data Science Catalyst workshop

**DOI:** 10.64898/2026.08.03.26359232

**Authors:** Katherine G Young, Amitava Banerjee, Colin Dayan, Spiros Denaxas, Sophie V Eastwood, Annie Jeffery, Martin K Rutter, Naveed Sattar, Jonathan Valabhji, Richard Horswood, Robin Humphreys, Michael Molete, Kay Murray, Peter Rogers, Dawn Veiro, Henry Ireland, Chrissie Walker, Beverley M Shields, Ewan Pearson, Andrew P McGovern, John M Dennis

**Author notes:** Corresponding author: Katherine G Young.

## Abstract

**Aims:** To develop a ‘core’ dataset of diabetes related variables to support reproducible research using UK routinely collected health data.

**Methods:** A workshop was conducted bringing together diabetes healthcare professionals, researchers, and patient and public representatives to discuss and prioritise variables for inclusion in the Diabetes Core Dataset. Core variables were those considered to be highest priority for diabetes research and available at high quality in NHS data routinely used for research (primary care [GP] and Hospital Episode Statistics [HES] data). Candidate variables for inclusion in the Diabetes Core Dataset were from a review of existing core datasets and expert opinion. Participants scored variables anonymously based on priority for diabetes research.

**Results:** 25 variables from existing diabetes core datasets and 87 other candidate variables were considered for inclusion in the Diabetes Core Dataset. All 25 of those from existing diabetes core datasets and 5 of the 87 candidate variables met the core requirements for inclusion. In addition, 7 variables were identified as high priority but not included in the core dataset as they are not currently available in GP/HES data; these were labelled as ‘future high priority’ variables for diabetes research.

**Conclusions:** A new diabetes core dataset for UK EHR research has been developed using a consensus-based process. The core dataset is openly available and can be flexibly applied in UK EHR (https://healthdatagateway.org/en/tool/426), including in new NHS Research Secure Data Environment platforms, to enhance reproducible research to improve the clinical care of people with diabetes and associated conditions.

## INTRODUCTION

The UK’s NHS medical data, in the form of anonymised electronic health records (EHR), are a world-leading resource for research to improve clinical care and benefit people living with diabetes. The UK EHR research landscape is rapidly evolving[1], with national and sub-national secure data environments (SDEs) becoming increasingly adopted as secure platforms allowing EHR research at-scale[2]. By providing single points of data access and data “airlocks”, SDE platforms ensure only accredited researchers can access health records, and prevent data leakage[3].

SDEs may also offer the opportunity to improve research quality[4]. If standardised disease phenotypes can be made available to researchers within the SDE infrastructure, this has the potential to save huge amounts of time for individual researchers. Standardisation can help reduce research waste, speed up research processes, and minimise inconsistencies between different studies due to variation in the underlying phenotype definitions.

Defining diabetes and its associated features for research using coded electronic health records is challenging and requires input from both diabetes healthcare professionals and data scientists. Major challenges include accurate identification of diabetes type[5] and date of disease onset; challenges directly addressed with the development of standardised algorithms in recent work by the Diabetes UK and British Heart Foundation funded Diabetes Data Science Catalyst (DDSC)[6]. However, beyond these elements a broader phenotypic library (“core dataset”) for diabetes research in UK EHR is not currently available, with existing datasets developed to support clinical care and audit rather than research[7].

We aimed to address the gap by developing an open-source core diabetes dataset: a minimum standardised set of variables required to characterise people with diabetes for most research applications, to support reproducible research using UK EHR. To achieve this, we first reviewed existing diabetes core datasets to identify priority diabetes-related variables, before convening a community of diabetes healthcare professionals, researchers, and patient and public representatives to discuss and agree the core dataset. We also aimed to identify important variables for diabetes research which are not currently available in the UK EHR datasets commonly used for research, with the aim of highlighting priorities for future data linkage beyond routine NHS data flows.

## METHODS

### Workshop participants

Healthcare professionals, researchers, and patient and public representatives were invited, via the Health Data Research (HDR) UK Diabetes Data Science Catalyst, to a two-hour online workshop held on 14^th^ November 2025. Seventeen participants attended (Appendix A); however, not all participated in every vote.

### Patient and public representatives

This workshop was conducted as part of an NHS Diabetes Driver project in collaboration with the NHS South West Secure Data Environment (SWSDE). The project involved regular fortnightly meetings with three patient and public representatives, including one of the SWSDE’s ‘Digital Critical Friends’, who provided ongoing input into project activities over the project duration (October 2025 to March 2026). In addition, three further patient and public representatives invited by HDR UK attended the workshop.

### Scope of the Diabetes Core Dataset

Prior to the workshop, we developed a definition for ‘core’ variables as those a) considered essential for most diabetes research projects (‘high priority’) and b) available at high quality in routinely accessible NHS research datasets (specifically primary care and secondary care Hospital Episode Statistics [HES] data). These two criteria were selected to ensure that ‘core’ variables can be made available across most NHS research platforms including the developing SDE network.

Patient identifiers required for data linkage and information describing the data source (for example, start and end of follow-up dates defined by patient registration dates in primary care) were considered structural requirements of the dataset and not discussed in the workshop.

For each core variable, we recommend that the dataset produced includes all available instances for each patient (i.e. a longitudinal structure) such as repeated BMI measurements or all recorded cardiovascular disease events, to maximise flexibility for downstream research use.

### Identification of pre-specified core variables

We conducted a pragmatic rapid literature review to identify existing diabetes core datasets described in peer-reviewed publications and grey literature. We searched PubMed/MEDLINE and conducted targeted web-based searches of relevant diabetes organisations, registries and audit programmes for diabetes core datasets, minimum datasets, and data standards. Searches combined “diabetes” with terms relating to data standardisation (“core dataset”, “minimum dataset”, “common data elements”, “standardised variables”, “data standards”). Searches were conducted in September 2025. We excluded datasets focused solely on specific interventions, outcomes, or individual studies without a broader data standardisation objective.

The review identified five pre-existing diabetes core datasets: two developed using NHS data primarily to support clinical care and audit (NHS Diabetes Summary Core Data Set[8], Core National Diabetes Audit[7]), and three designed for broader research and clinical trial applications across different data sources and settings (SCORE-IT[9], EUBIROD[10], German Center for Diabetes Research [DZD] Core Data Set[11]). The Scottish Diabetes Core Dataset[12], also developed using NHS data to support clinical care, was additionally included following recommendation from a study team member with expertise in diabetes data standards. The PRSB Diabetes Information Standard[13], recently developed by the NHS to produce a standard for sharing diabetes information between people and professionals, was not considered a ‘core’ diabetes dataset as it contains many variables which are not currently available in routine NHS data, including those directly from digital technology and medical devices.

Variables present in at least three of the six existing core datasets were included in a list of 25 ‘pre-specified core variables’ which were assumed to meet both core requirements (high priority and available in routine NHS research data). For each variable, data sources and their underlying domains (e.g. demographics, diabetes features, clinical measurements) were extracted to allow a comparison of included variables across datasets (Supplementary Table 1). Some variables were combined into broader categories to allow alignment between the datasets (e.g. different facets of cardiovascular disease combined into a single cardiovascular disease variable group).

### Identification of potential additional core variables and future priority variables

87 additional variables potentially relevant to diabetes research were identified through the literature review and expert opinion (‘candidate variables’). During the workshop, participants were also invited to contribute additional variables to this list.

The additional variable list was intentionally not restricted to those available in routine NHS research data, as the workshop also aimed to determine ‘future high priority variables’ beyond the core dataset to inform priorities for future data linkage outside routine NHS data flows. These variables were defined as those considered important for diabetes research by participants, but not yet routinely available in current NHS-based research datasets.

### Workshop structure

During the first phase of the workshop, participants were presented with the list of 25 ‘pre-specified core variables’ identified from existing core diabetes datasets (Supplementary Table 2) and asked to vote on whether any should be excluded from the Diabetes Core Dataset (Figure 1) based on perceived lack of importance for diabetes research.

**Figure 1:**
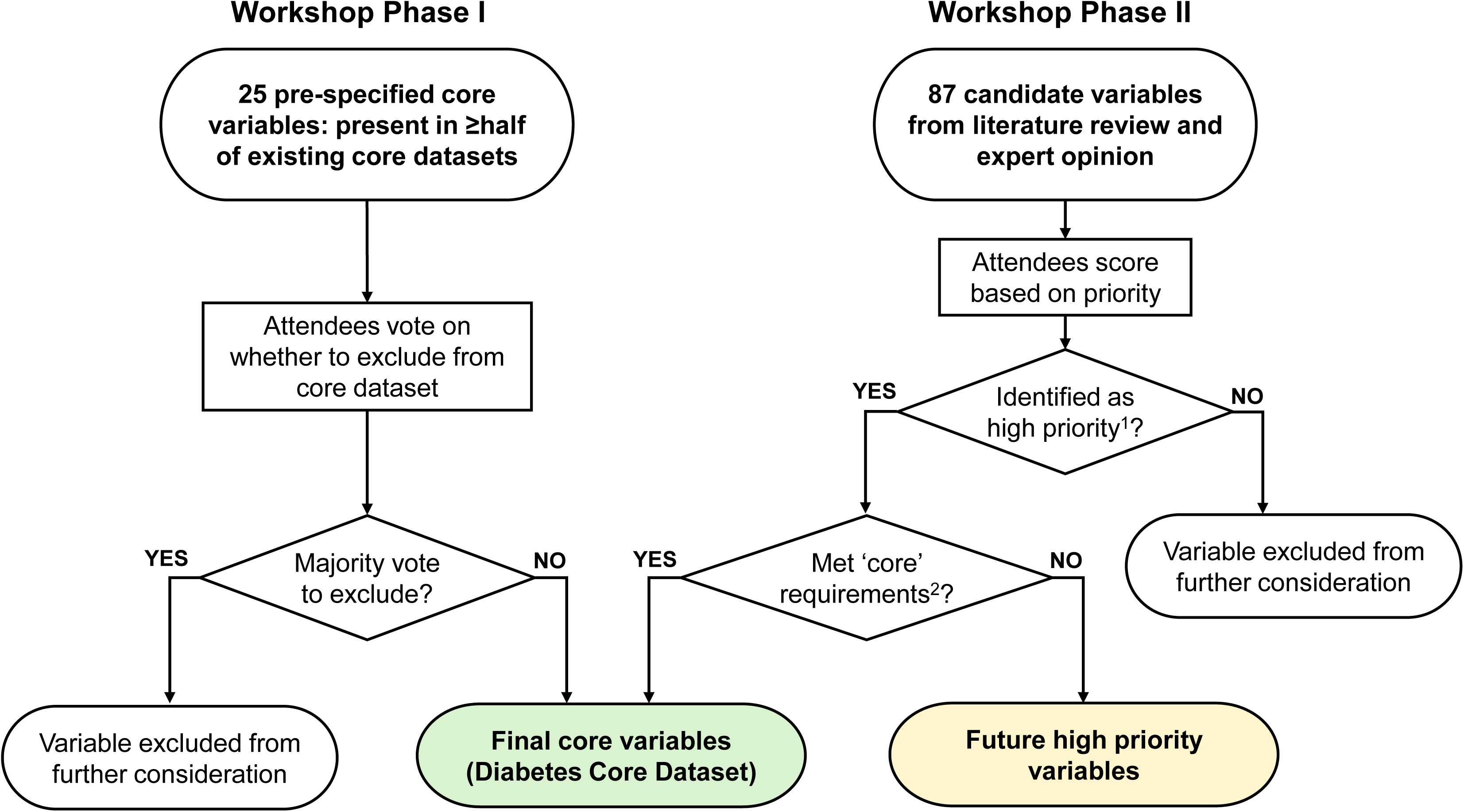
Workshop structure. Participant input was captured in two phases: in Phase 1 participants were presented with a list of 25 ‘pre-specified’ core variables identified from pre-existing diabetes core datasets and asked to vote on whether to exclude these from the Diabetes Core Dataset. In Phase 2, participants were presented with 87 candidate variables and asked to score these on a 1-5 scale based on importance for diabetes research. ^1^High priority was defined as top 50% based on researcher score or top 25% based on PPI score. ^2^The requirements for ‘core’ variables were being high priority for diabetes research and available at high quality in GP/HES data; the latter was decided by authors Young, McGovern and Dennis. Non-diabetes conditions which are not common diabetes complications, or medications for these conditions, were also excluded from the Diabetes Core Dataset as these were considered more appropriately addressed by disease-specific core datasets.

During the second phase, participants were asked to score each of the 87 ‘candidate variables’ on a 5-point scale, based on how important they felt the variable was for diabetes research (Figure 1). Participants were also able to contribute additional variables not already included, which were then made available for scoring in real-time. Participants were asked to score based on priority only, ignoring whether variables met the ‘core’ definition, in order to understand what variables are important but not currently available for research.

All voting was anonymous, but participants had the option to identify as either a ‘researcher’ (including clinicians) or ‘PPI member’ (patient/public representative).

### Consensus process

Following the first phase of the workshop, variables from the pre-specified list were excluded from the Diabetes Core Dataset if the majority of participants voted for exclusion.

Following the second phase of the workshop, a pragmatic threshold was used to define high vs low priority after reviewing the distribution of scores, with the aim of retaining variables of comparable importance to diabetes research as those in the pre-specified dataset. ‘High priority’ variables were defined as those in the top 50% of researcher scores or in the top 25% of PPI member scores. This approach placed greater emphasis on researcher-derived prioritisation, reflecting their greater familiarity with data requirements for research, while ensuring that variables highly valued by PPI contributors were also retained.

High priority variables were then assessed by authors Young, McGovern and Dennis to determine whether they met the second ‘core’ requirement: availability at high quality in NHS GP and HES data. High priority variables meeting this requirement were included in the Diabetes Core Dataset. Medical conditions which are not considered common microvascular or macrovascular complications of diabetes were not included in the Diabetes Core Dataset as we felt they would be better addressed by other core datasets developed by experts in relevant specific disease areas.

High priority variables that did not meet ‘core’ requirements due to their lack of availability in NHS GP and HES data were classified as ‘future high priority variables’. These variables were separated into variables specific to diabetes research, including diabetes focused treatments, processes of care or complications, and those non-specific to diabetes research.

## RESULTS

Voting during the first phase of the workshop, involving 15 participants (5 patient/public representatives and 10 researchers), led to all 25 pre-specified core variables being included in the Diabetes Core Dataset. Full voting results are reported in Supplementary Table 2.

In the second phase of the workshop, participants scored 87 candidate variables based on their priority for diabetes research (Full voting results: Supplementary Table 3). 50 variables were identified as high priority (Supplementary Table 4). 45 of these were in the top 50% of researcher scores, and an additional 5 were included which were in the top 25% of the patient/public representative scores but not in the top 50% of researcher scores: anti-depressant use, genetic ancestry, diabetes symptoms at diagnosis, highest education level, and treatment for severe mental illness.

Of the 50 high priority variables, five were considered to be available at high quality in routine NHS research data (primary care and secondary care Hospital Episode Statistics [HES] data) and therefore met the requirements to be included in the Diabetes Core Dataset. These variables were: liver function tests (Alanine Aminotransferase levels [ALT] only were included in the core dataset as other tests were not routinely available), and four treatment categories: lipid lowering, blood-pressure lowering, anti-platelet and prescriptions for continuous glucose monitoring (CGM) devices. As a result, the final Diabetes Core Dataset comprised a total of 30 core variables (Figure 2).

**Figure 2:**
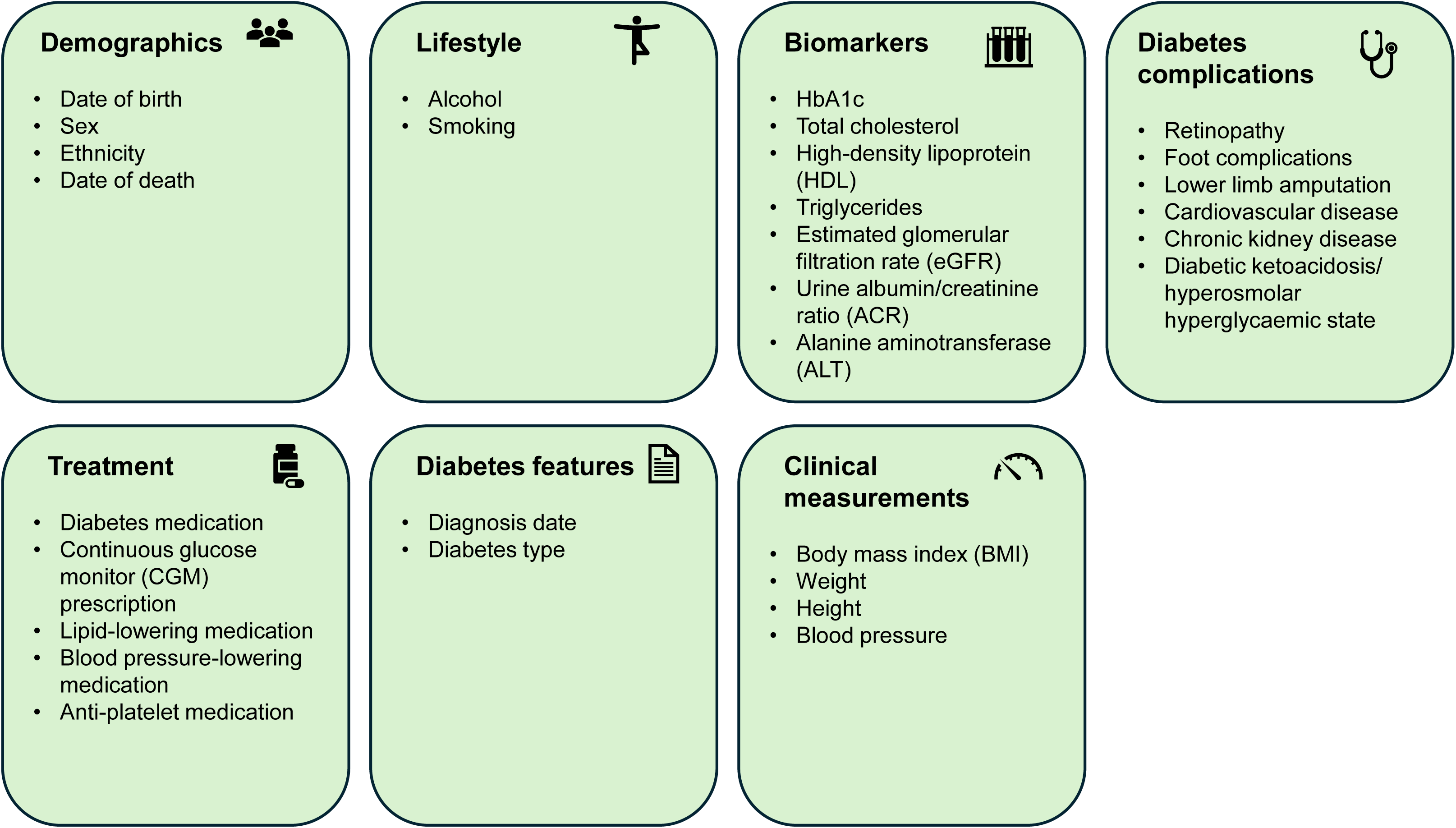
Variables in the final Diabetes Core Dataset. We recommend that all available instances of each variable are extracted for each patient (i.e. a longitudinal structure) such as repeated BMI measurements or all recorded cardiovascular disease events, to maximise flexibility for downstream research use.

Additionally, 18 ‘future high priority variables’ were identified as they met core requirements but are available only in alternative data flows to NHS GP and HES data. 7 of these were classified as having specific relevance to diabetes research: high quality graded retinopathy data (e.g. grading performed using the UK National Screening Committee (NSC) grading classification), continuous glucose monitoring results data, results of genetic testing for monogenic forms of diabetes (MODY), insulin dose/timing information from insulin pumps and smart pens, private prescriptions for GLP-1 analogues and related medications, data on hypoglycaemia requiring emergency care, and use of community podiatry services for foot complications. Table 1 further specifies these variables as well as potential data feeds that could capture these variables.

**Table 1:** Future high priority data items for diabetes research from alternative data feeds beyond GP and HES data. These are ordered by researcher priority (highest priority at the top of the table).

| Domain | Data item | Potential additional data feed(s) |
| --- | --- | --- |
| Diabetes complications | High quality graded retinopathy data (e.g. 'left eye M1R1, right eye M0R0') | Grade data from Diabetic Eye Screening Programme (DESP) |
| Biomarkers | Continuous glucose monitoring (CGM) results | CGM data from patients or commercial providers |
| Omics | MODY (Maturity-Onset Diabetes of the Young) genetic testing results | Detailed secondary care data / data from NHS Genomic Medicine Service |
| Treatment of diabetes and associated conditions | Insulin pump/smart pen data on insulin dose/timing | Insulin data from patients |
| Treatment of diabetes and associated conditions | Private prescriptions for incretin-based therapies | Private healthcare provider prescribing records, and community and private pharmacy dispensing data |
| Diabetes complications | Hypoglycaemia requiring emergency care | Ambulance and Accident & Emergency data |
| Diabetes complications | Use of community podiatry services (foot complications) | Community podiatry service data |

Supplementary Table 5 reports the 11 remaining high priority data items not specific to diabetes research and available through additional data feeds. These included demographic data (census-level ethnicity data, social deprivation, cause of death), health resources utilisation, granular data on cardiovascular disease (e.g. ejection fraction), environmental variables (walkability, rural-urban location, density of fast food outlets), private healthcare usage, and pregnancy history.

## DISCUSSION

We have developed an expert- and patient-led consensus core dataset for diabetes and associated features in UK EHR. The dataset contains 30 core variables, and can be applied across UK EHR where primary and secondary care data are available. Key variables include demographic and lifestyle factors, diabetes type, date of diagnosis, clinical measurements, biomarkers, and common diabetes complications. To support research reproducibility and uptake, the core dataset is publicly accessible at: https://exeter-diabetes.github.io/sde-sw-framework/ and via the HDR UK Health Data Research Gateway (https://healthdatagateway.org/en/tool/426).

Key strengths of this work are that the Diabetes Core Dataset is publicly available and easily implementable. The dataset is designed around data fields available for most people with diabetes within routinely accessible NHS research data sources. It integrates existing algorithms for diabetes phenotyping in UK EHRs, such as a recent algorithm for classifying diabetes type developed through the Diabetes Data Science Catalyst and validated using genetic data[6]. The intended purpose is for the core dataset to provide a flexible yet standardised starting dataset providing diabetes and related phenotypes relevant for most research studies. This starting dataset can then be extended through curation of study specific variables and/or data linkages to answer specific research questions of interest.

Implementation of the core dataset could reduce duplication of effort by providing a shared foundation for diabetes research and enabling use by researchers without specialist diabetes expertise. It may also facilitate more consistent incorporation of diabetes into wider epidemiological and clinical research, whether as an exposure, covariate, or outcome. Adoption of the core dataset can therefore both reduce research waste and improve research reproducibility and quality.

The identification of ‘future high priority variables’ for diabetes research beyond the core dataset, including retinopathy, CGM, and ambulance data, and potential data flows to access them, represents an additional key output of the workshop. These high priority variables were systematically derived from participant rankings of perceived importance for diabetes research, incorporating both researcher and patient and public views. Patient/public representatives highlighted the importance of variables such as mental health, symptoms at diagnosis, and education level, which were subsequently prioritised despite being ranked less highly by researchers. A key aim of the NHS Research SDE network is to facilitate novel data linkages, and the future high priority variable list we have identified both highlights key current gaps in routine NHS research data flows, and could inform a systematic approach to prioritising future novel data linkages to maximise patient benefit. Limitations of the core dataset include the use of a pragmatic consensus-based approach to identify and prioritise variables, which, while transparent and reproducible, may not capture all perspectives or reflect a formally structured consensus methodology (e.g. Delphi process). The thresholds used to define high priority variables were also set pragmatically following review of score distributions, which may introduce some subjectivity into classification.

By design, the Diabetes Core Dataset represents a minimum dataset intended to provide a common foundation for diabetes related research. As such, it does not attempt to encompass either the broad range of acute and chronic conditions experienced by people living with diabetes, or by extension the full breadth of variables that may be required for a typical research study in practice (e.g. risk factors for diabetes. The dataset was also primarily developed with type 1 and type 2 diabetes in mind, reflecting their predominance in routine clinical care and research. Consequently, variables of particular importance for rarer forms of diabetes may not be included.

Defining the codelists and algorithms required to derive all core variables from EHR data was beyond the scope of this workshop. While we highlight the availability of the recent consensus algorithm to classify diabetes type[6], equivalent validated phenotyping definitions for diabetes complications including cardiovascular conditions were more limited. In the absence of these, publicly available repositories including the HDR UK Phenotype Library[14] and OpenCodelists[15] provide an array of open source codelists and phenotyping algorithms encompassing all variables included in the core dataset. Future work is needed to develop and maintain more standardised phenotype definitions following FAIR (Findable, Accessible, Interoperable, and Reusable) principles.

Finally, any deployment of the dataset remains dependent on the quality and completeness of underlying routine healthcare data. Issues such as coding variation (regional and temporal), missingness, variation in laboratory measurements and reporting practices, and misclassification (including potential misclassification of diabetes type, duration, and complications) are likely to persist and may introduce bias in downstream analyses despite standardisation of variable selection.

### Future work

Our immediate next stages of development include active roll-out and testing of the core dataset across the regional SDE network. This will assess the feasibility of implementing standardised variable definitions across different data environments, identify data quality and availability issues, and refine extraction approaches to support reproducible generation of the dataset. In the future, this may enable federated research using a national-scale Diabetes Core Dataset. Such an approach aligns with the ambitions of the emerging Health Data Research Service by enabling secure access to data at national scale, while maintaining local curation expertise, governance processes, and public trust, alongside the ability to provide bespoke datasets[16].

In parallel, we will work to enable access to future priority data flows which could be integrated into the core dataset. Future developments could also include expansion of the existing core dataset to more broadly include pre-diabetes and related conditions, such as obesity and wider cardiometabolic disease.

Beyond diabetes, the approach we have developed to define the Diabetes Core Dataset provides a template for the pragmatic, low-cost development of disease-specific core datasets across other clinical areas, incorporating input from health professionals, researchers, people living with the disease, and the public. Many variables are likely to be common across multiple disease areas, allowing shared components to be reused with disease-specific features incorporated where required. Such core datasets may also have potential to improve routine data quality by informing clinical recording templates and quality improvement initiatives, for example through incorporation into primary care systems.

Adoption of this model of expert-developed, disease-specific core datasets could rapidly increase the capacity of the SDE network to support high quality and novel research using UK EHR.

### Conclusions

A new diabetes core dataset for UK EHR research has been developed using a transparent consensus-based process with input from health professionals, researchers, patients and the public. The core dataset is openly available and can be flexibly applied in UK EHR, including in the new national and sub-national SDE network, to enhance reproducible research to improve the clinical care of people with diabetes and associated conditions.

## Data Availability

All data produced in the present study are shown in full in the Supplementary Materials.

## Acknowledgements

We thank all participants in the Diabetes Data Science Catalyst workshop, including healthcare professionals, researchers, and patient and public contributors, for their time and input into the development of the Diabetes Core Dataset.

We acknowledge the support of Health Data Research UK and the Diabetes Data Science Catalyst in facilitating workshop delivery, and the NIHR Exeter Biomedical Research Centre for supporting this work.

## Funding information

This work was supported as part of a driver project funded through the NHS Data for Research and Development Programme, delivered via the NHS Research Secure Data Environment (SDE) Network and funded by NHS England. Additional support was provided by the Diabetes Data Science Catalyst, funded by Diabetes UK and the British Heart Foundation in collaboration with Health Data Research UK.

SD is supported by: a) BHF Data Science Centre / CVD-COVID-UK/COVID-IMPACT consortium, led by HDR UK (SP/19/3/34678), b) NIHR Biomedical Research Centre at University College London Hospital NHS Trust (UCLH BRC), c) a BHF Accelerator Award (AA/18/6/24223), d) Multimorbidity Mechanism and Therapeutic Research Collaborative (MMTRC, grant number MR/V033867/1), e) NIHR-UKRI CONVALESCENCE study, and the Longitudinal Health and Wellbeing COVID-19 National Core Study, which was established by the UK Chief Scientific Officer in October, 2020, and funded by UKRI (grant references MC_PC_20030 and MC_PC_20059). SVE is funded by a British Heart Foundation UCL Centre for Research Excellence Springboard Fellowship. JMD is supported by a Wellcome Trust Early-Career Award (227070/Z/23/Z).

The funders had no role in study design, data collection, analysis, interpretation, or manuscript preparation. The views expressed are those of the authors and not necessarily those of the NIHR, NHS or the Department of Health and Social Care.

## Conflict of Interest Statement

MKR reports consultancy payments from Eli Lilly, unrelated to this work. NS has consulted for and/or received speaker honoraria from AbbVie, Amgen, AstraZeneca, Boehringer Ingelheim, Carmot Therapeutics, Eli Lilly, Gan & Lee, GlaxoSmithKline, Hanmi Pharmaceuticals, Kailera, Mass Medicines, Menarini-Ricerche, Metsera, Novo Nordisk, Pfizer, Regeneron, Roche, UCB Pharma, and Verdiva Bio; and received grant support paid to his University from AstraZeneca, Boehringer Ingelheim, Novartis, and Roche outside the submitted work. JV was the National Clinical Director for Diabetes and Obesity at NHS England from 2013 to 2023. No other authors report potential conflicts of interest.

## Appendix A: Workshop participants

Amitava Banerjee

Colin Dayan

Spiros Denaxas

John Dennis

Sophie Eastwood

Annie Jeffery

Ewan Pearson

Martin Rutter

Naveed Sattar

Jonathan Valabhji

*Anonymous*

Patient/public representatives:

Richard Horswood

Robin Humphreys

Mike Molete

Kay Murray

Peter Rogers

Dawn Veiro

