## Supplementary Material for "A consensus diabetes core dataset for research using NHS data: outputs from a Diabetes Data Science Catalyst workshop"

### Supplementary Data

| Domain | No. | Data item | NHS Diabetes Summary Core Data Set [1] | Core National Diabetes Audit [2] | Scottish Diabetes Core Dataset [3] | SCORE-IT [4] | EUBIROD [5] | DZD Core Data Set [6] | Total |
|---|---|---|---|---|---|---|---|---|---|
| Demographics and social determinants of health | 1 | DOB | X | X | X | | X | X | 5 |
| | 2 | Sex | X | X | X | | X | X | 5 |
| | 3 | Ethnicity | | X | X | | | X | 3 |
| | 4 | Date of death | X | | X | X | | | 3 |
| Diabetes features | 5 | Age/date of diabetes diagnosis | X | X | X | | X | X | 5 |
| | 6 | Diabetes type (including remission) | X | X | X | | X | X | 5 |
| Clinical measurements | 7 | BMI | X | X | X | | X | | 4 |
| | 8 | Weight | | X | X | X | X | X | 5 |
| | 9 | Height | | X | X | | X | X | 4 |
| | 10 | Blood pressure | X | X | X | | X | X | 5 |
| Biomarkers | 11 | HbA1c | X | X | X | X | X | X | 6 |
| | 12 | Lipids: total cholesterol | X | X | X | | X | X | 5 |
| | 13 | Lipids: HDL | | | X | | X | X | 3 |
| | 14 | Lipids: triglycerides | | | X | | X | X | 3 |
| | 15 | Kidney function: eGFR | X | X | X | X | X | X | 6 |
| | 16 | Kidney function: urine ACR | X | X | X | X | X | X | 6 |
| Diabetes complications | 17 | Retinopathy | X | | X | X | X | X | 5 |
| | 18 | Foot complications (ulcer, infection, loss of sensation/pulse) | | X | X | X | X | | 4 |
| | 19 | Lower limb amputation (minor and major) | X | | X | X | X | X | 5 |
| | 20 | Cardiovascular disease: overall and by subtype (including hypertension and heart failure) | X | X | X | X | X | X | 6 |
| | 21 | Chronic kidney disease stage plus transplant, dialysis | X | | X | X | X | X | 5 |

| | | | | | | | | | |
|---|---|---|---|---|---|---|---|---|---|
| | 22 | Diabetic ketoacidosis (DKA)/ hyperosmolar hyperglycaemic state (HHS) | X | | X | X | | | 3 |
| Lifestyle | 23 | Alcohol status | | | X | | X | X | 3 |
| | 24 | Smoking status | X | X | X | | X | X | 5 |
| Diabetes treatment | 25 | Diabetes glucose-lowering medication | | X | X | | X | X | 4 |

**Supplementary Table 1: Sources for candidate core variables.** Candidate variables were harmonised across source datasets where appropriate. Broad constructs (e.g., glycaemic control in SCORE-IT) were mapped to specific routinely collected measures (HbA1c), and related concepts (e.g., different aspects of cardiovascular disease) were rationalised into common variable definitions.

| Domain | No. | Data item | Researcher votes to exclude | PPI votes to exclude |
|---|---|---|---|---|
| Demographics and social determinants of health | 1 | DOB | 0/10 | 0/5 |
| | 2 | Sex | 0/10 | 0/5 |
| | 3 | Ethnicity | 0/10 | 0/5 |
| | 4 | Date of death | 0/10 | 2/5 |
| Diabetes features | 5 | Age/date of diabetes diagnosis | 0/10 | 0/5 |
| | 6 | Diabetes type (including remission) | 0/10 | 0/5 |
| Clinical measurements | 7 | BMI | 0/10 | 1/5 |
| | 8 | Weight | 0/10 | 0/5 |
| | 9 | Height | 1/10 | 0/5 |
| | 10 | Blood pressure | 0/10 | 0/5 |
| Biomarkers | 11 | HbA1c | 0/10 | 0/5 |
| | 12 | Lipids: total cholesterol | 0/10 | 0/5 |
| | 13 | Lipids: HDL | 0/10 | 0/5 |
| | 14 | Lipids: triglycerides | 0/10 | 0/5 |
| | 15 | Kidney function: eGFR | 0/10 | 0/5 |
| | 16 | Kidney function: urine ACR | 0/10 | 0/5 |
| Diabetes complications | 17 | Retinopathy | 0/10 | 0/5 |
| | 18 | Foot complications (ulcer, infection, loss of sensation/pulse) | 0/10 | 0/5 |
| | 19 | Lower limb amputation (minor and major) | 0/10 | 0/5 |
| | 20 | Cardiovascular disease: overall and by subtype (including hypertension and heart failure) | 0/10 | 0/5 |
| | 21 | Chronic kidney disease stage plus transplant, dialysis | 0/10 | 0/5 |
| | 22 | Diabetic ketoacidosis (DKA)/ hyperosmolar hyperglycaemic state (HHS) | 0/10 | 0/5 |
| Lifestyle | 23 | Alcohol status | 0/10 | 0/5 |
| | 24 | Smoking status | 0/10 | 0/5 |
| Diabetes treatment | 25 | Diabetes glucose-lowering medication | 0/10 | 0/5 |

**Supplementary Table 2: Voting results from Phase I of the workshop.** 10 out of 11 researcher participants and 5 out of 6 PPI participants voted on whether to exclude any of the 25 pre-specified variables from the Diabetes Core Dataset.

| Domain | No. | Data item | From atten-dees | Researcher mean score | Researcher vote count | Researcher score rank | PPI mean score | PPI vote count | PPI score rank |
|---|---|---|---|---|---|---|---|---|---|
| Demographics and social determinants of health | 1 | Accommodation (ownership, number in household, care home, park home) | 0 | 3 | 10 | 73 | 2.83 | 6 | 72 |
| | 2 | Cause of death | 0 | 4.91 | 11 | 1 | 3.33 | 6 | 44 |
| | 3 | Census-level ethnicity data | 1 | 4.57 | 7 | 8 | 2.6 | 5 | 79 |
| | 4 | Census-level family structure | 1 | 3.71 | 7 | 55 | 3.25 | 4 | 48 |
| | 5 | Deprivation (area-level: IMD/Townsend) | 0 | 4.55 | 11 | 11 | 3.83 | 6 | 17 |
| | 6 | Education (highest level) | 0 | 3.64 | 11 | 60 | 3.83 | 6 | 17 |
| | 7 | Employment (full/part-time, retired, student, type of work) | 0 | 3.73 | 11 | 54 | 2.67 | 6 | 78 |
| | 8 | Family history of diabetes | 0 | 4.64 | 11 | 7 | 4.5 | 6 | 2 |
| | 9 | Genetic ancestry | 0 | 3.7 | 10 | 58 | 4.17 | 6 | 9 |
| | 10 | Household income | 0 | 3.82 | 11 | 48 | 3.8 | 5 | 24 |
| | 11 | Pet ownership | 1 | 1.67 | 6 | 87 | 2.25 | 4 | 84 |
| Environment | 12 | Density of fast-food outlets | 0 | 4.18 | 11 | 32 | 2.83 | 6 | 72 |
| | 13 | Density of gambling establishments | 0 | 2.91 | 11 | 78 | 2 | 5 | 87 |
| | 14 | Distance to GP | 0 | 3.82 | 11 | 48 | 3.5 | 6 | 33 |
| | 15 | Green space/coastal proximity | 0 | 3.55 | 11 | 64 | 2.5 | 6 | 81 |
| | 16 | Noise | 0 | 2.55 | 11 | 84 | 2.17 | 6 | 85 |
| | 17 | Pollution | 0 | 3 | 11 | 73 | 2.17 | 6 | 85 |
| | 18 | Rural-urban (area-level) | 0 | 4 | 11 | 37 | 2.5 | 6 | 81 |
| | 19 | Walkability | 0 | 4.27 | 11 | 26 | 3 | 6 | 58 |
| Diabetes features | 20 | Autoantibodies | 0 | 4.4 | 10 | 17 | 4 | 6 | 12 |
| | 21 | C-peptide | 0 | 4.36 | 11 | 21 | 4.17 | 6 | 9 |
| | 22 | Glucose at diagnosis | 0 | 3.91 | 11 | 41 | 4.17 | 6 | 9 |
| | 23 | Referral to diabetes self-management program (DAFNE etc.) | 1 | 3.86 | 7 | 45 | 3 | 5 | 58 |
| | 24 | Symptoms at diagnosis | 0 | 3.7 | 10 | 58 | 4 | 6 | 12 |
| Clinical measurements | 25 | Heart rate | 0 | 3 | 10 | 73 | 3.2 | 5 | 51 |
| | 26 | Hip circumference | 0 | 4 | 11 | 37 | 3.33 | 6 | 44 |
| | 27 | Thigh circumference (proxy of muscle mass) | 1 | 2.33 | 6 | 85 | 2.8 | 5 | 76 |
| | 28 | Waist circumference | 0 | 4.45 | 11 | 13 | 3.83 | 6 | 17 |
| Biomarkers | 29 | Blood urea | 0 | 2.67 | 9 | 81 | 2.83 | 6 | 72 |
| | 30 | C-reactive protein | 0 | 3.44 | 9 | 66 | 3.6 | 5 | 31 |
| | 31 | CGM results | 0 | 4.4 | 10 | 17 | 3.5 | 6 | 33 |
| | 32 | Fasting glucose | 0 | 2.89 | 9 | 79 | 3.5 | 6 | 33 |
| | 33 | Full blood count (WBC, RBC, haematocrit, Hb, MCH, MCHC, MCV, platelets) | 0 | 3.11 | 9 | 71 | 3 | 5 | 58 |
| | 34 | Further urine tests e.g. WBC, RBC, nitrite, pH | 0 | 1.89 | 9 | 86 | 3 | 5 | 58 |
| | 35 | LDL | 0 | 3.44 | 9 | 66 | 2.83 | 6 | 72 |
| | 36 | Liver function (AST, ALT, albumin) | 0 | 4.67 | 9 | 6 | 3.83 | 6 | 17 |

| | | | | | | | | | |
|---|---|---|---|---|---|---|---|---|---|
| | 37 | NT-proBNP | 1 | 3.8 | 5 | 50 | 3.33 | 3 | 44 |
| | 38 | Sodium/potassium | 0 | 2.78 | 9 | 80 | 3 | 6 | 58 |
| | 39 | Thyroid stimulating hormone | 0 | 3.11 | 9 | 71 | 3.17 | 6 | 54 |
| Diabetes complications | 40 | High quality retinopathy grading | 0 | 4.78 | 9 | 5 | 3.5 | 6 | 33 |
| | 41 | Hypoglycaemia requiring emergency care | 0 | 4.22 | 9 | 31 | 4.67 | 6 | 1 |
| | 42 | More granular cardiovascular data e.g. LVEF | 0 | 4.44 | 9 | 14 | 3.67 | 6 | 25 |
| | 43 | Use of community podiatry services (foot complications) | 0 | 3.89 | 9 | 42 | 3.67 | 6 | 25 |
| Comorbidities | 44 | Autoimmune conditions | 0 | 4.11 | 9 | 36 | 4.5 | 6 | 2 |
| | 45 | Cancer | 0 | 4.56 | 9 | 9 | 3.17 | 6 | 54 |
| | 46 | Dementia | 0 | 4.44 | 9 | 14 | 4.2 | 5 | 7 |
| | 47 | ECHO data | 1 | 3.5 | 6 | 65 | 3 | 3 | 58 |
| | 48 | Family history of (premature) ASCVD | 1 | 3.86 | 7 | 45 | 2.8 | 5 | 76 |
| | 49 | Frailty | 0 | 4.33 | 9 | 22 | 3.67 | 6 | 25 |
| | 50 | GI illnesses e.g. GERD | 1 | 3 | 5 | 73 | 3.67 | 3 | 25 |
| | 51 | Gum disease | 1 | 2.6 | 5 | 82 | 3 | 4 | 58 |
| | 52 | History of emergency hospitalisations | 1 | 3.86 | 7 | 45 | 3 | 6 | 58 |
| | 53 | Impotence | 0 | 4.12 | 8 | 35 | 3.4 | 5 | 42 |
| | 54 | Learning disability | 0 | 4.44 | 9 | 14 | 3.2 | 5 | 51 |
| | 55 | Liver disease | 0 | 4.56 | 9 | 9 | 4.33 | 6 | 4 |
| | 56 | Mental illness | 0 | 4.33 | 9 | 22 | 3.83 | 6 | 17 |
| | 57 | Musculoskeletal diseases | 1 | 2.6 | 5 | 82 | 3.5 | 4 | 33 |
| | 58 | Respiratory disease | 0 | 3.22 | 9 | 70 | 3.33 | 6 | 44 |
| | 59 | Sleep apnoea | 1 | 4.4 | 5 | 17 | 3.25 | 4 | 48 |
| | 60 | Vascular ultrasound | 1 | 3.33 | 6 | 69 | 3 | 5 | 58 |
| Lifestyle | 61 | Diet | 0 | 4.14 | 7 | 33 | 3.5 | 6 | 33 |
| | 62 | Physical activity | 0 | 4.29 | 7 | 24 | 4.33 | 6 | 4 |
| | 63 | Received dietary advice from healthcare professional | 0 | 3.71 | 7 | 55 | 3 | 5 | 58 |
| | 64 | Sleep | 0 | 4 | 7 | 37 | 3.17 | 6 | 54 |
| | 65 | Weight management programmes | 0 | 4.25 | 8 | 27 | 4.2 | 5 | 7 |
| Treatment of diabetes and associated conditions | 66 | Anti-platelet therapy | 0 | 4.38 | 8 | 20 | 3 | 5 | 58 |
| | 67 | Blood-pressure lowering medication | 0 | 4.88 | 8 | 2 | 4 | 6 | 12 |
| | 68 | CGM prescription | 1 | 4.83 | 6 | 4 | 3.25 | 4 | 48 |
| | 69 | Health resource utilisation | 0 | 4.5 | 8 | 12 | 2.4 | 5 | 83 |
| | 70 | Insulin pump/smart pen data | 0 | 4.25 | 8 | 27 | 3.83 | 6 | 17 |
| | 71 | Lipid lowering medication | 0 | 4.88 | 8 | 2 | 3.6 | 5 | 31 |
| | 72 | Private healthcare usage | 0 | 3.88 | 8 | 43 | 3.4 | 5 | 42 |
| | 73 | Private prescriptions for GLP1 | 1 | 4.25 | 4 | 27 | 3.67 | 3 | 25 |
| | 74 | Side effects from diabetes medications | 0 | 3.62 | 8 | 61 | 3.5 | 6 | 33 |

| | | | | | | | | | |
|---|---|---|---|---|---|---|---|---|---|
| | 75 | Whether diabetes managed in primary or secondary care | 0 | 3.75 | 8 | 52 | 3 | 6 | 58 |
| | 76 | Whether self-monitoring blood glucose | 0 | 4 | 7 | 37 | 3.67 | 6 | 25 |
| Other treatments | 77 | Anti-depressant use | 1 | 3.8 | 5 | 50 | 4 | 3 | 12 |
| | 78 | Oral steroid use | 1 | 4.25 | 4 | 27 | 3.5 | 4 | 33 |
| | 79 | Treatment for severe mental illness - antipsychotics | 1 | 3.6 | 5 | 62 | 4.33 | 3 | 4 |
| Reproductive health | 80 | Details of pregnancies including number of children | 0 | 3.88 | 8 | 43 | 3.83 | 6 | 17 |
| | 81 | Menopause status/age at menopause | 0 | 3.75 | 8 | 52 | 3.5 | 6 | 33 |
| Omics | 82 | MODY test results | 0 | 4.29 | 7 | 24 | 3 | 6 | 58 |
| | 83 | Proteomics | 0 | 3 | 7 | 73 | 2.6 | 5 | 79 |
| | 84 | T1D and T2D polygenic risk scores | 0 | 3.71 | 7 | 55 | 3.2 | 5 | 51 |
| | 85 | Whole genome sequencing | 0 | 3.43 | 7 | 68 | 3.17 | 6 | 54 |
| Patient-reported outcome measures | 86 | Diabetes distress questionnaires and similar | 0 | 3.57 | 7 | 63 | 3 | 5 | 58 |
| | 87 | Quality of Life / activities of daily living | 0 | 4.14 | 7 | 33 | 4 | 6 | 12 |

**Supplementary Table 3: Scores from Phase II of the workshop.** Table shows the numbers of researchers (‘researcher vote count’) and PPI representative (‘PPI vote count’) who scored each variable. Each participant scored each variable on a 1-5 scale based on priority for diabetes research (1=lowest priority, 5=highest priority). Mean scores grouped by participant type are shown, as well as score rank (1=highest ranked by researchers/participants). ‘From attendees’ indicates variables suggested by attendees during the workshop.

| Domain | No. | Data item | Researcher score rank | PPI score rank | Meet core requirements? | Reason for not meeting requirements |
|---|---|---|---|---|---|---|
| Demographics and social determinants of health | 2 | Cause of death | 1 | 44 | No | Requires additional data feed(s) |
| Treatment of diabetes and associated conditions | 67 | Blood-pressure lowering medication | 2 | 12 | Yes | |
| Treatment of diabetes and associated conditions | 71 | Lipid lowering medication | 2 | 31 | Yes | |
| Treatment of diabetes and associated conditions | 68 | CGM prescription | 4 | 48 | Yes | |
| Diabetes complications | 40 | High quality retinopathy grading | 5 | 33 | No | Requires additional data feed(s) |
| Biomarkers | 36 | Liver function (AST, ALT, albumin) | 6 | 17 | Yes | ALT only meets core requirements of being available at high quality in GP/HES data |
| Demographics and social determinants of health | 8 | Family history of diabetes | 7 | 2 | No | Not well captured in any routine data source |
| Demographics and social determinants of health | 3 | Census-level ethnicity data | 8 | 79 | No | Requires additional data feed(s) |
| Comorbidities | 45 | Cancer | 9 | 54 | No | Better addressed by disease-specific core dataset |
| Comorbidities | 55 | Liver disease | 9 | 4 | No | Better addressed by disease-specific core dataset |
| Demographics and social determinants of health | 5 | Deprivation (area-level: IMD/Townsend) | 11 | 17 | No | Requires additional data feed(s) |
| Treatment of diabetes and associated conditions | 69 | Health resource utilisation | 12 | 83 | No | Requires additional data feed(s) |
| Clinical measurements | 28 | Waist circumference | 13 | 17 | No | Not well captured in any routine data source |
| Diabetes complications | 42 | More granular cardiovascular data e.g. LVEF | 14 | 25 | No | Requires additional data feed(s) |
| Comorbidities | 46 | Dementia | 14 | 7 | No | Better addressed by disease-specific core dataset |
| Comorbidities | 54 | Learning disability | 14 | 51 | No | Better addressed by disease-specific core dataset |
| Diabetes features | 20 | Autoantibodies | 17 | 12 | No | Not well captured in any routine data source |

| | | | | | | |
|---|---|---|---|---|---|---|
| Biomarkers | 31 | CGM results | 17 | 33 | No | Requires additional data feed(s) |
| Comorbidities | 59 | Sleep apnoea | 17 | 48 | No | Better addressed by disease-specific core dataset |
| Treatment of diabetes and associated conditions | 66 | Anti-platelet therapy | 20 | 58 | Yes | |
| Diabetes features | 21 | C-peptide | 21 | 9 | No | Not well captured in any routine data source |
| Comorbidities | 49 | Frailty | 22 | 25 | No | Better addressed by disease-specific core dataset |
| Comorbidities | 56 | Mental illness | 22 | 17 | No | Better addressed by disease-specific core dataset |
| Lifestyle | 62 | Physical activity | 24 | 4 | No | Not well captured in any routine data source |
| Omics | 82 | MODY test results | 24 | 58 | No | Requires additional data feed(s) |
| Environment | 19 | Walkability | 26 | 58 | No | Requires additional data feed(s) |
| Lifestyle | 65 | Weight management programmes | 27 | 7 | No | Requires additional data feed(s) |
| Treatment of diabetes and associated conditions | 70 | Insulin pump/smart pen data | 27 | 17 | No | Requires additional data feed(s) |
| Treatment of diabetes and associated conditions | 73 | Private prescriptions for GLP1 | 27 | 25 | No | Requires additional data feed(s) |
| Other treatments | 78 | Oral steroid use | 27 | 33 | No | Better addressed by disease-specific core dataset |
| Diabetes complications | 41 | Hypoglycaemia requiring emergency care | 31 | 1 | No | Requires additional data feed(s) |
| Environment | 12 | Density of fast-food outlets | 32 | 72 | No | Requires additional data feed(s) |
| Lifestyle | 61 | Diet | 33 | 33 | No | Not well captured in any routine data source |
| PROMs and related | 87 | Quality of Life / activities of daily living | 33 | 12 | No | Not well captured in any routine data source |
| Comorbidities | 53 | Impotence | 35 | 42 | No | Better addressed by disease-specific core dataset |
| Comorbidities | 44 | Autoimmune conditions | 36 | 2 | No | Better addressed by disease-specific core dataset |
| Environment | 18 | Rural-urban (area-level) | 37 | 81 | No | Requires additional data feed(s) |

| | | | | | | |
|---|---|---|---|---|---|---|
| Clinical measurements | 26 | Hip circumference | 37 | 44 | No | Not well captured in any routine data source |
| Lifestyle | 64 | Sleep | 37 | 54 | No | Not well captured in any routine data source |
| Treatment of diabetes and associated conditions | 76 | Whether self-monitoring blood glucose | 37 | 25 | No | Not well captured in any routine data source |
| Diabetes features | 22 | Glucose at diagnosis | 41 | 9 | No | Not well captured in any routine data source |
| Diabetes complications | 43 | Use of community podiatry services (foot complications) | 42 | 25 | No | Requires additional data feed(s) |
| Treatment of diabetes and associated conditions | 72 | Private healthcare usage | 43 | 42 | No | Requires additional data feed(s) |
| Reproductive health | 80 | Details of pregnancies including number of children | 43 | 17 | No | Requires additional data feed(s) |
| Other treatments | 77 | Anti-depressant use | 50 | 12 | No | Better addressed by disease-specific core dataset |
| Demographics and social determinants of health | 9 | Genetic ancestry | 58 | 9 | No | Not well captured in any routine data source |
| Diabetes features | 24 | Symptoms at diagnosis | 58 | 12 | No | Not well captured in any routine data source |
| Demographics and social determinants of health | 6 | Education (highest level) | 60 | 17 | No | Not well captured in any routine data source |
| Other treatments | 79 | Treatment for severe mental illness - antipsychotics | 62 | 4 | No | Better addressed by disease-specific core dataset |

**Supplementary Table 4: Results from Phase II of the workshop: high priority variables ranked by researcher score.** High priority variables (top 50% of researcher scores or top 25% of PPI scores) were assessed by authors Young, McGovern and Dennis for whether they fulfilled the second core requirement of being available at high quality in NHS data routinely used for research (GP/HES data). Those which fulfilled this requirement were included in the Diabetes Core Dataset, except for non-diabetes conditions which are not common diabetes complications, or medication for these conditions, as these were considered more appropriately addressed by disease-specific core datasets. Those which require additional feeds were identified as 'future high priority' variables and included in Table 1 (diabetes-specific variables) and Supplementary Table 5 (non-diabetes-specific variables).

| Domain | Data item | Potential additional data feed(s) |
|---|---|---|
| Demographics and social determinants of health | Cause of death | Office for National Statistics death data |
| Demographics and social determinants of health | Census-level ethnicity data | Census data |
| Demographics and social determinants of health | Deprivation (area-level: IMD/Townsend) | UK government area-level deprivation measures (data is readily available but requires secure linkage to patient postcode) |
| Treatment of diabetes and associated conditions | Health resource utilisation | Data from all health services |
| Diabetes complications | More granular cardiovascular data e.g. LVEF | Detailed secondary care data |
| Environment | Walkability | Various datasets available including from Ordnance Survey and Office for National Statistics |
| Lifestyle | Weight management programmes | Data from weight management programmes |
| Environment | Density of fast-food outlets | Public Health England area-level data |
| Environment | Rural-urban (area-level) | Office for National Statistics Rural-Urban area-level classification |
| Treatment of diabetes and associated conditions | Private healthcare usage | Data from private healthcare providers |
| Reproductive health | Details of pregnancies including number of children | Data from secondary care maternity services |

**Supplementary Table 5: Future high priority data items, not specific to diabetes research, from alternative data feeds beyond GP and HES data.**
